# Clinical Prediction Models for Treatment Outcomes in Newly-diagnosed Epilepsy

**DOI:** 10.1101/2024.01.12.24301215

**Authors:** Corey Ratcliffe, Vishnav Pradeep, Anthony Marson, Simon S. Keller, Laura J. Bonnett

## Abstract

**Background:** Up to 35% of individuals diagnosed with epilepsy proceed to develop pharmacoresistant epilepsy, leading to persistent uncontrolled seizure activity that can directly, or indirectly, significantly degrade an individual’s quality of life. The factors underlying pharmacoresistance are unclear, but it has been hypothesised that repeated ictogenic activity is conducive to the development of a more robust epileptogenic network. To ensure that the most effective treatment choices are made and ictogenic activity is minimised, accurate outcome modelling at the point of diagnosis is key.

**Objectives:** This review therefore aims to identify demographic, clinical, physiological (e.g. EEG), and imaging (e.g. MRI) factors that may be predictive of treatment outcomes in patients with newly diagnosed epilepsy (NDE).

**Data sources, study eligibility criteria, participants, and interventions:** MEDLINE and EMBASE were searched for prediction models of treatment outcomes in patients with newly diagnosed epilepsy and any non-surgical treatment plan.

**Study appraisal and synthesis methods:** Study characteristics were extracted and subjected to assessment of risk of bias (and applicability concerns) using the PROBAST tool. Prognostic factors associated with treatment outcomes are reported.

**Results:** After screening, 48 models were identified in 32 studies, which generally scored low for concerns of applicability, but universally high for susceptibility to bias. Outcomes reported were heterogenous, but fit broadly into four categories: pharmacoresistance, short-term treatment response, seizure remission, and mortality. Prognostic factors were also heterogenous, but the predictors that were commonly significantly associated with outcomes were those related to seizure characteristics (semiology), epilepsy history, and age at onset. ASM response was often included as a prognostic factor, potentially obscuring factor relationships at baseline.

**Conclusions:** Currently, outcome prediction models for NDE demonstrate a high risk of bias. Model development could be improved with a stronger adherence to recommended TRIPOD practices, and by avoiding including response to treatment as a prognostic factor.

**Implications of key findings:** This review identified semiology, epilepsy history, and age at onset as factors associated with treatment outcome prognosis, suggesting that future prediction model studies should focus on these factors in their models. Furthermore, we outline actionable changes to common practices that are intended to improve the overall quality of prediction model development in NDE.

**Key Points:**

- **This paper presents a systematic literature search for treatment outcome prediction models in newly diagnosed epilepsy.**
- **The risk of bias in the included models were evaluated using the PROBAST framework, finding a universally high risk level.**
- **The relationship between semiology, epilepsy history, and age at onset with seizure remission should be examined in future prediction model studies.**
- **To improve the overall quality of prediction model development in NDE, prospective authors are advised to adhere to TRIPOD guidelines, and to avoid including response to treatment as a prognostic variable.**

## Introduction

### Rationale

#### Clinical overview of epilepsy

As one of the most common neurological diseases, epilepsy is estimated to affect over 70 million people globally.^1,2^ Epilepsy incidence tends to be higher in the youngest and oldest age groups, in females, and in low-middle income countries.^3^ Epilepsy is characterised by a predisposition to spontaneous seizure activity, which is thought to arise due to abnormalities within cortical networks.^4–7^ Epilepsy may refer to any subtype of a multitude of disorders that can differ in seizure type, clinical course, or prognosis; aetiology and comorbidities will also inform the specific diagnoses people with epilepsy (PWE) will receive.^8,9^ An accurate diagnosis is crucial for determining the appropriate first-line treatment, which will most commonly be anti-seizure medication (ASM) monotherapy. Alongside seizure activity, PWE are vulnerable to cognitive, behavioural, and neurological comorbidities, the combination of which often result in PWE experiencing a lower quality of life.

#### Newly-diagnosed epilepsy

To better characterise the course of epilepsy and its underlying pathomechanisms, it has been suggested that (people with) newly diagnosed epilepsy (NDE) be studied as a distinct group.^10,11^ Studying epilepsy at its earliest time point avoids the confounds inherent in long-standing epilepsy, including the chronic effects of seizure activity and ASM use: seizure activity in chronic epilepsy can cause injuries that encourage the development of pharmacoresistance in PWE, and successive ASM regimens are associated with a reduction in the chance of attaining seizure freedom.^12–15^ Predictive models for chronic epilepsy consequently include the confounding variability of seizure and ASM profiles, whereas models developed specifically for use in NDE (which can reliably prognosticate disease trajectories) do not. Models based on NDE cohorts can therefore inform treatment approaches that ameliorate the consequences of epilepsy chronicity.

#### Seizure freedom and pharmacoresistance in First-line Therapies

In a recent large-cohort NDE study, the rate of 1-year remission (cessation of seizure activity) following ASM mono/polytherapy was 63.7%, and the rate of pharmacoresistance (failure of two or more appropriate ASM trials to control seizure activity) was 36.3%, in line with similar studies.^16–18^ ASM considered efficacious for focal seizure management include oxcarbazepine, carbamazepine, and lamotrigine; for generalised seizures, sodium valproate, levetiracetam, lamotrigine, and ethosuximide are common first-line ASM.^19–22^ First-line monotherapy represents the best likelihood of preventing seizure recurrence and pharmacoresistance in PWE who have had two seizures or been classified as ‘high-risk’, and an optimal treatment response is most likely when the appropriate ASM is chosen for monotherapy.^23–26^ If a change of ASM is indicated, it is recommended that alternative monotherapy be prescribed.^27^ Should this prove unsuccessful, polytherapy, adjunctive therapy, or surgical therapies (i.e. two or more ASM, dietary management, and resection respectively) may be considered.^20^

### Second-line Therapies

The success of polytherapy is largely dependent on the choice of ASM combination. Combination ASM are often determined based on complementary mechanisms of action, but no specific selection criteria exist, due in part to difficulty in collecting randomised control trial (RCT) data for combination approaches.^18^ Should ASM therapy fail, the ketogenic diet is recommended as an adjunctive therapy, and has been found to be effective in encouraging seizure control in people with pharmacoresistant epilepsy, as have invasive techniques of seizure control in focal epilepsies, such as resective surgery, vagus nerve stimulation, deep brain stimulation, and responsive neuromodulation.^28,29^ It has been suggested that 12 months after diagnosis is the ideal time at which to evaluate the long-term efficacy of treatment, especially considering the implications for driving restrictions in Europe.^30,31^ Furthermore, 12 months is the earliest possible timepoint at which a PWE can be said to have experienced 12 months of continuous seizure freedom, and a referral for surgical therapy is also unlikely.^32,33^

### Treatment Outcomes

Early seizure control has been indicated to be crucial for ensuring optimal treatment outcomes in NDE, putatively due to the prevention of further disruptions to seizure-related networks.^5,34–37^ Epilepsy treatment is individualised to ensure that: a) The risk-benefit ratio of a proposed therapy is suitable and b) The PWE is receiving the most efficacious treatment.^27^ The decision to begin a particular regimen is made after the consideration of several potential contraindications, such as pregnancy, medical interactions, and the risk of adverse effects.^38^ Importantly, the treatment choice will also be informed by the likelihood of achieving seizure freedom on a particular ASM (the efficacy) and the proportion of PWE who persist with the drug trial (the effectiveness). Predicting treatment outcomes—such as seizure remission, refractoriness, and pharmacoresistance—is non-trivial, also requiring the consideration of factors like age at onset (and the related epilepsy duration), the number of pretreatment seizures, EEG/imaging abnormalities, intellectual impairments, aetiology, and semiology to inform trajectories.^39–42^

### Prediction models

Prediction models are combinations of prognostic factors used to estimate the risk of a specific endpoint. Built with and validated on large cohorts, prediction models allow for individual patient outcomes to be estimated according to a formal statistical framework.^43^ Prognostic and diagnostic models are commonplace in epilepsy care, and the principal benefit of multivariable models (over the use of univariable factors for prediction) is accuracy, especially considering the complexity of epileptic processes.^44,45^ Single biomarkers (quantifiable properties indicative of normal biological processes) in epilepsy are thought to lack the granularity and robustness necessary to allow for clinical application.^46^ For example, several studies have investigated the relationship between EEG abnormalities and outcomes, often providing conflicting or incongruous evidence; whilst it is probable that some association exists, it is likely that EEG patterns and features influence/are mediated by external factors, and further multivariable research is required to determine how.^47^ To facilitate application and future evaluation (as in with systematic reviews) it is recommended that prediction models be designed and reported in a systematic manner, such as is outlined by Transparent Reporting of a multivariable prediction model for Individual Prognosis Or Diagnosis (TRIPOD) guidelines.^48^ Adherence to a predefined set of guidelines, such as TRIPOD, helps to ensure that the risk of bias (RoB; systematic error) and applicability concerns (AC) in the resultant study are kept to a minimum.^49^ Several models for the prediction of treatment outcomes in NDE have already been proposed, the latest systematic review of which was published in 2014.^50,51^

### Objectives

RCTs are the gold-standard source for evidence of efficacy and effectiveness for therapies, but it has previously been remarked that there is a lack of suitable high-quality evidence in the literature, with many of the existing studies suffering from methodological flaws.^52^ Alternatively, systematic reviews (of prediction models, for example) provide a reliable collation of evidence from which health-based interventions can be informed, without contributing to research waste.^53,54^ As new prediction models are developed and validated, it is crucial that they be presented in a format that allows for optimal dissemination of their actionable conclusions. Furthermore, information from previous reviews may be outdated and misleading in the context of more recent findings. The most recent comparable review, carried out by Abimbola et al. in 2014, presents several opportunities for improvement (besides being updated), namely that only studies with samples of over 100 were included and no evaluation of RoB was carried out.^50^ A systematic examination of multivariable prediction models for treatment outcomes in NDE was undertaken here to provide an updated and expanded review of the state of the literature, and to facilitate understanding of their conclusions. All included models were evaluated for RoB and AC using the PROBAST (Prediction model Risk Of Bias ASsessment Tool) framework.^49^ Between the models, common prognostic factors were identified and are presented herein, with the intention of informing future prediction model studies in NDE.

## Methods

### Protocol and registration

This review is reported in adherence with the PRISMA (Preferred Reporting Items for Systematic reviews and Meta-Analyses) guidelines, and a non-peer reviewed, publicly available, protocol was registered with PROSPERO (ID: CRD42022329936).^55^

### Eligibility Criteria, Information Sources, Search, Study Selection, and Data Collection Process

MEDLINE and Embase were searched for relevant publications, using PubMed/MeSH (medical subject headings) and Scopus/Boolean terms respectively. Full queries can be seen in Appendix 1. Data were screened by CR and LJB independently, with mediation of any conflicting exclusions following consensus meetings provided by SSK. Studies were included if they contained a multivariable model of treatment outcomes in a discrete sample of NDE, meeting the following criteria:

- **Study design - Any primary design including (but not limited to): cohort studies, randomised control trials, quasi-randomised control trials, observational studies, and case-control studies.**
- **Participants - Any person with NDE defined using the operational ILAE definition of two clinically unprovoked seizures, or one unprovoked seizure with a > 60% probability of recurrence (other definitions were evaluated for agreement with the ILAE definition *ad hoc*).^56^ Provoked seizures include those deemed situational or due to acute neurological insult/precipitant.^57^**
- **Multivariable model - Prediction models developed with at least two demographic, and/or clinical, and/or neuroimaging, and/or electrophysiological factors collected and assessed as part of standard clinical practice at baseline upon a new diagnosis of epilepsy, that are associated with 12 months continuous seizure freedom (remission). Demographic factors are socioeconomic attributes that can be statistically expressed—for example age, sex, and education level. Clinical factors are signs and symptoms of disease classification or severity including aetiology, type and frequency of seizure, age of onset of epilepsy and duration of illness prior to diagnosis. The neuroimaging and neurophysiological factors include assessments of standard MRI and EEG examinations respectively, often taken upon a new diagnosis of epilepsy.**

– **Our search terms were not designed to capture machine learning/artificial intelligence-based studies, due to the complexities introduced by the structure of these models, which is very different to those of regression-based models.**^58^
- **Primary outcomes - 12 months (or longer) continuous seizure freedom (remission). The timeframe of 12 months was chosen in accordance with previous literature suggesting that as one seizure per year is sufficient to preclude PWE from driving, seizure freedom should be measured over the same timeframe.**^17^ **Furthermore, after 12 months of treatment, if seizures are not controlled it has been recommended that the PWE be referred to a specialist clinic.**^59^
- **Secondary outcomes - Reported seizure remission of any duration at any time point; treatment failure (adverse effects, intractability, etc.) reported in any form and at any time.**

### Risk of Bias in Individual Studies

RoB was determined on a per-study basis, using 20 signalling questions over four domains (Participants, Predictors, Outcomes, Analysis); the answers to the questions indicate potential for bias, which then informs the (semi-subjective) potential for bias in that domain. If any domain is flagged as having a high potential for bias, the study is judged to have a high overall RoB.^54^ Similarly, three of the four domains contain an AC judgement, whereby the rater evaluates to what extent the study content matches the research question. High concern for applicability in any domain results in the study also receiving a high AC rating.^49^ Alongside outcome-related data extraction, data required for RoB and AC assessment were extracted by CR and VP, who independently evaluated all 32 studies in the sample.

### Summary Measures, Synthesis of Results, Risk of Bias Across Studies, and Additional Analyses

Data pertinent to describing the setting, methodology, demographics, predictors, and outcomes for individual studies was synthesised into narrative form and evidence tables. Metadata for quality assessments purposes was also extracted. Sankey plots were constructed to visually present the distribution of outcomes across studies, and predictors across outcomes. Definitions for the categories proposed in this study can be found in Appendix 2.

## Results

### Study Selection

After the removal of 285 duplicate entries, 878 records were excluded first based on their titles, then abstracts. The remaining 128 reports were sought for retrieval, of which 126 were obtained. The retrieved reports were then assessed for eligibility, during which 77 were excluded due to univariable modelling (44), unsuitable cohorts (20), unsuitable outcomes (12), or absence of primary analysis (1). The remaining reports underwent data extraction, during which 17 were deemed ineligible.^60^ Data extraction was carried out on the whole sample by CR and VP independently, using a predefined form to ensure that all relevant information was extracted systematically.

### Study Characteristics

After screening, 32 studies were deemed suitable for inclusion (for a PRISMA diagram, see Figure 1), including 48 models. 12 studies used prospectively recruited PWE (37.5%), 17 used retrospective data (53.1%), and three used a combination (9.4%). Designs included one case-control study, two RCTs, and 29 cohort studies. Sample sizes ranged from 53 to 99990 PWE, with a median value of 261. Estimates of ‘events per variable’ ranged from 0.63 to 3927.38, with a median value of 9.86 (for study characteristics, see table 1). In our sample, 12 studies utilised Cox proportional hazards models (37.5%), whereas the remaining 20 employed logistic regressions (62.5%) to build their prediction models. Outcomes were evaluated at timepoints that ranged from 16 - 20 weeks, up to 32 - 36 years, with several studies assessing outcomes at the arbitrary date of the last follow-up.

**Figure 1.**
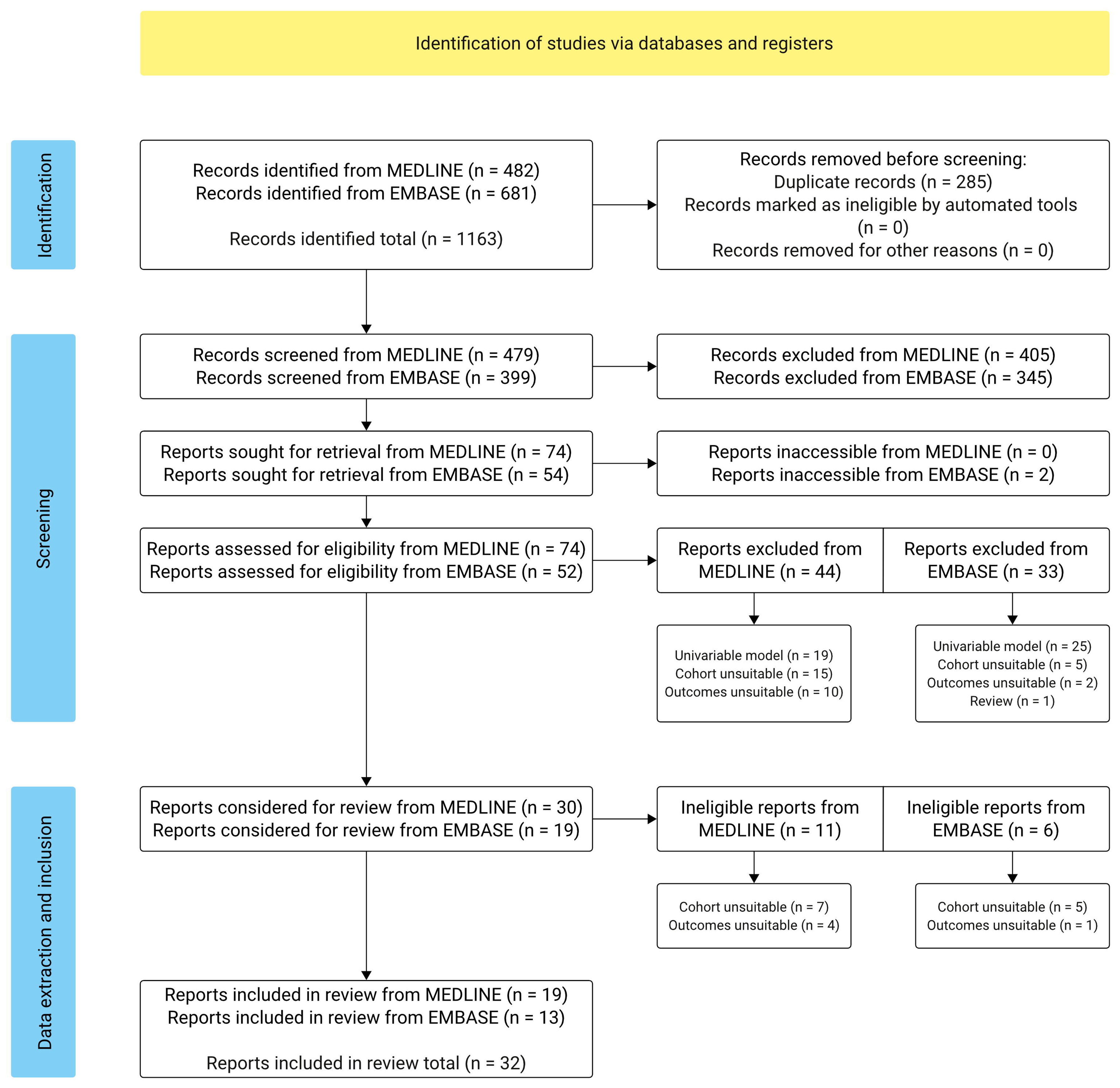
PRISMA flow diagram of study selection.

**Table 1.**
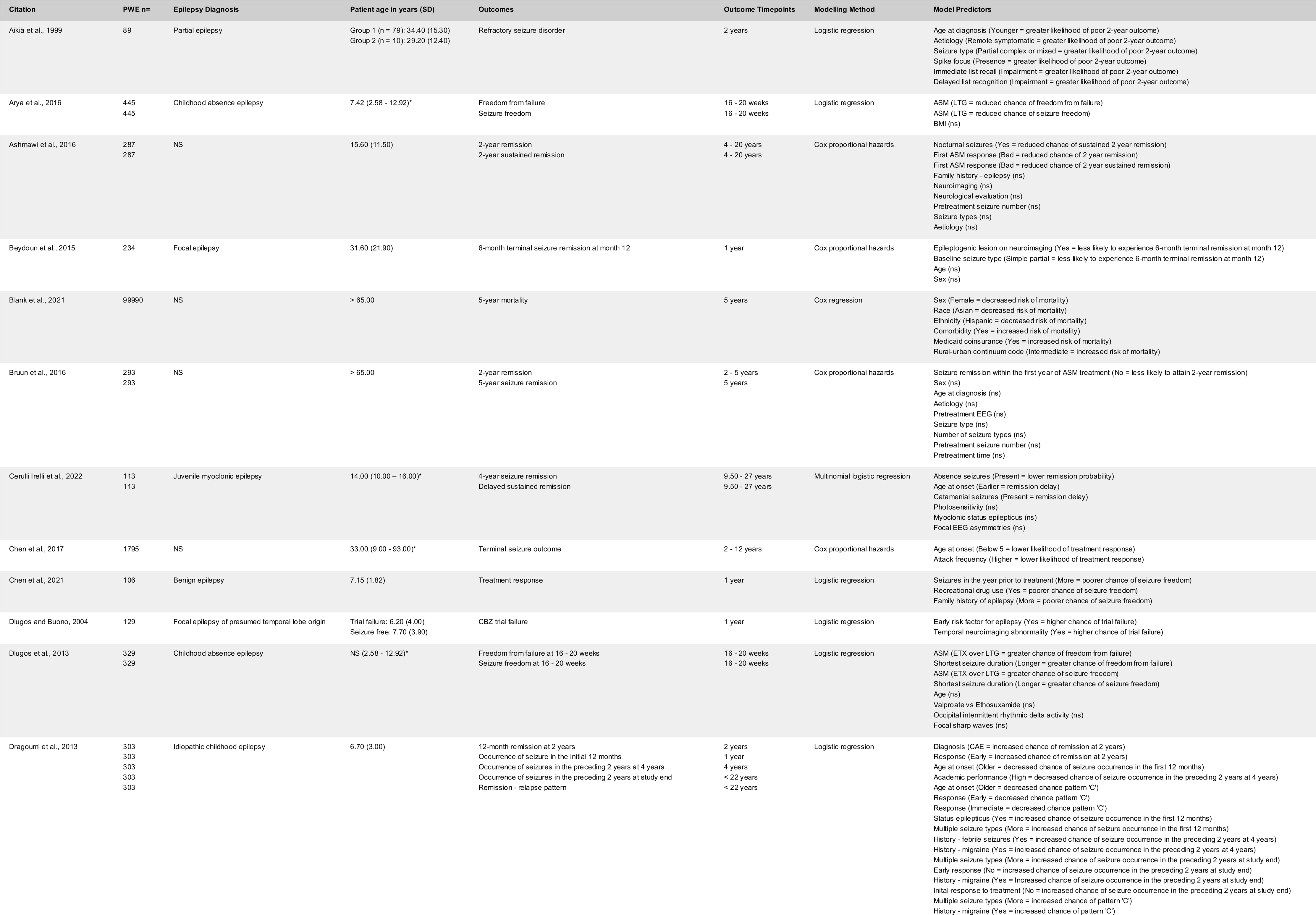
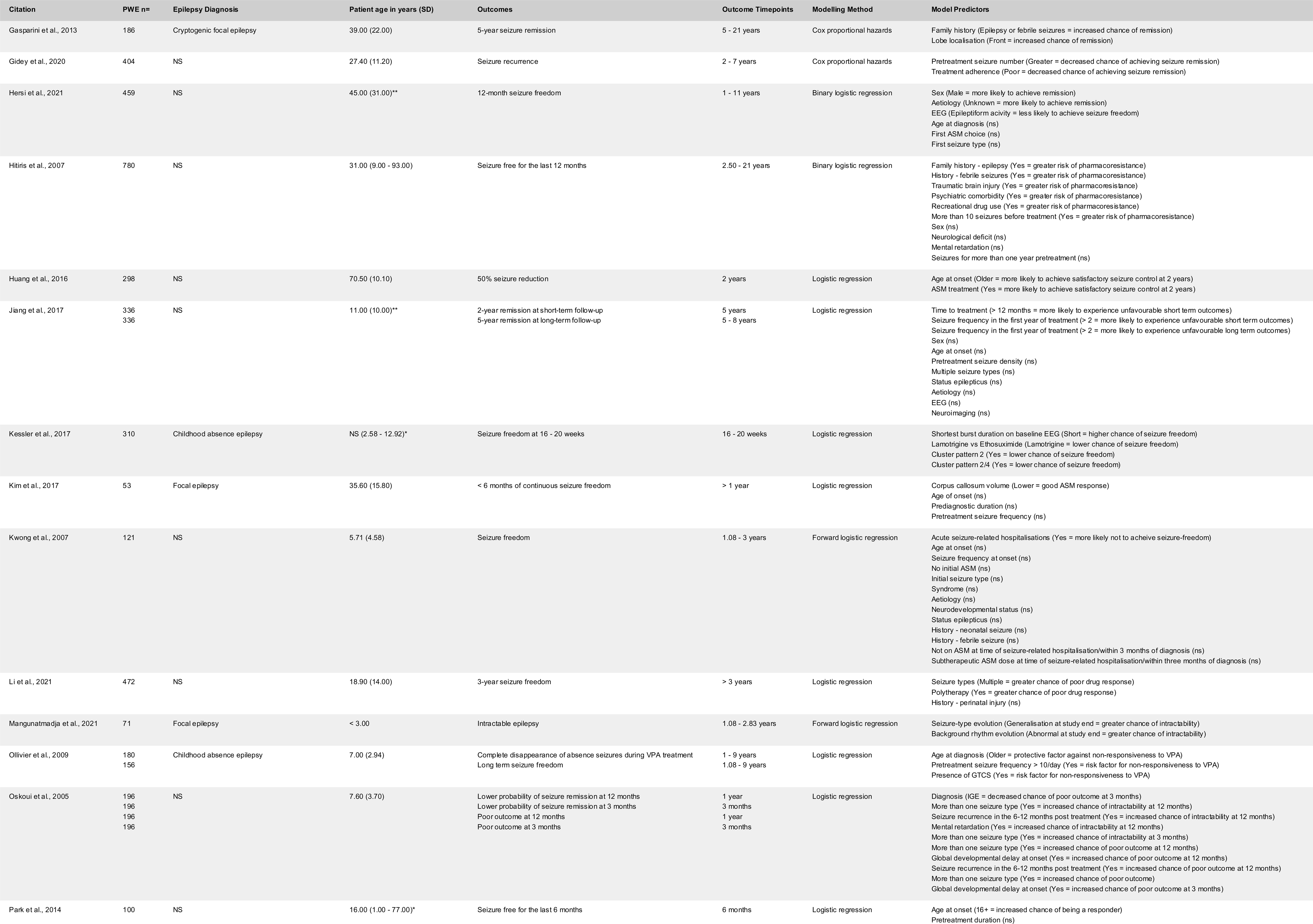
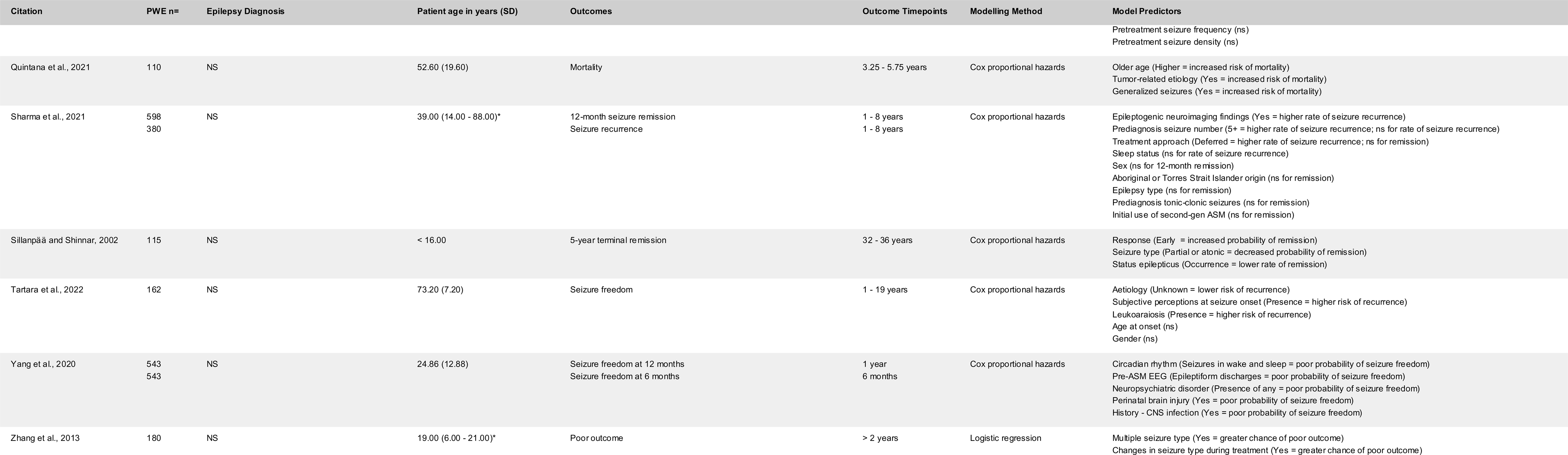
Summary of included studies. Where available, ages are presented in years as mean (SD), *Median (range), or **Median (IQR). Epilepsy diagnoses are reported as in the original studies.^16,39,42,61–89^

Across all models, 41 unique outcomes were operationalised, which were subsequently stratified into four categories (see figure 2): Mortality; Pharmacoresistance; Seizure remission; and Short-term treatment response (a complete list of outcomes is provided in Appendix 3). In accordance with the review objectives, the seizure remission category was used for seizure outcomes of 12 months or longer, with all seizure outcomes of less than 12 months being categorised as short-term treatment response. 69 unique predictors were operationalised, which were subsequently stratified into 11 categories: Age; ASM; Comorbidity; Demographics; Diagnosis; EEG; History; Neuroimaging; Neuropsychology; Response; and Semiology (a complete list of predictors is also provided in Appendix 3). Although unavailable at baseline, response (to treatment) variables were recorded in a number of studies, and contributed significant predictors to several multivariable prediction models.

**Figure 2.**
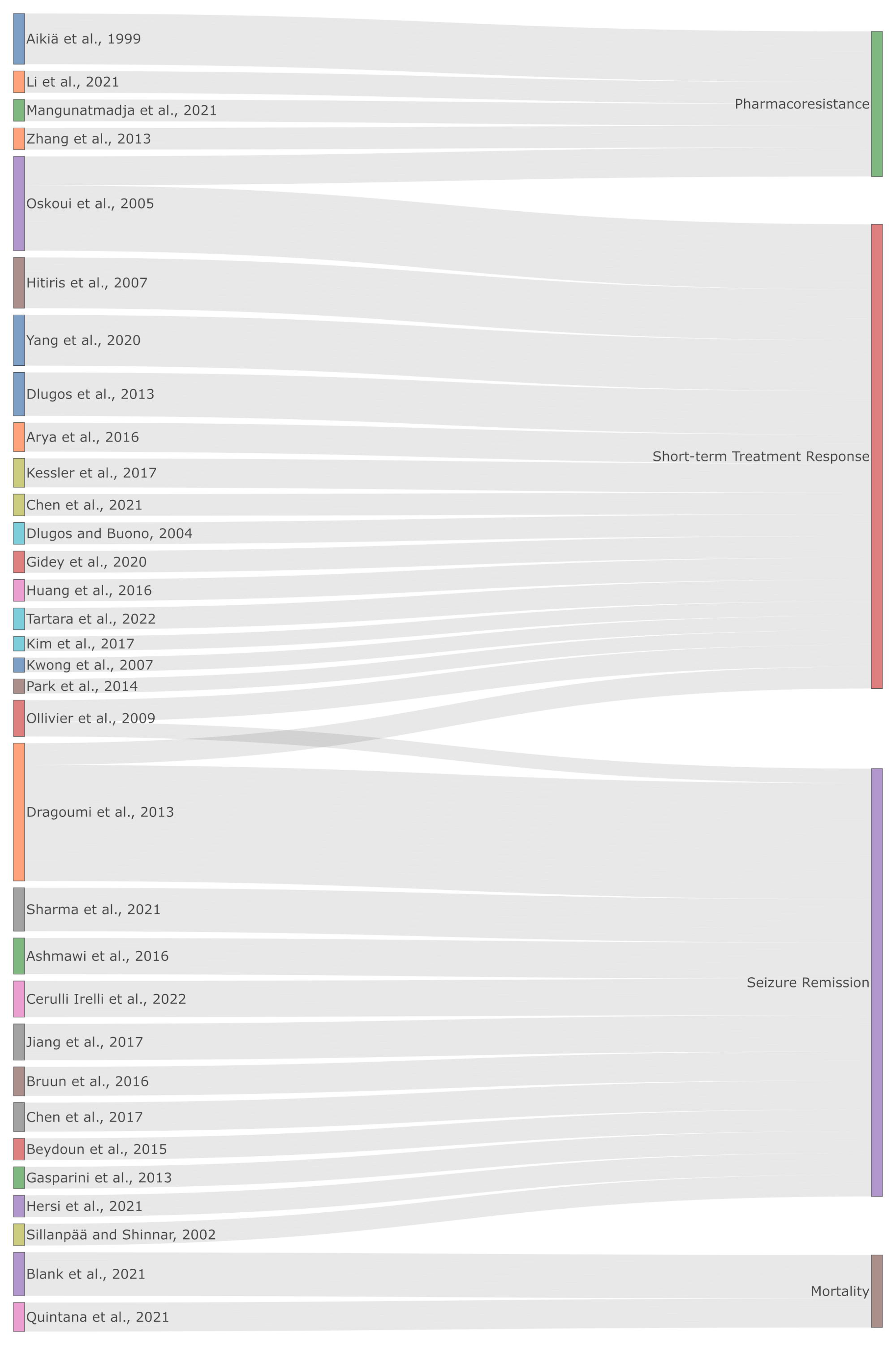
Sankey diagram showing model outcomes across all studies.^16,39,42,61–89^.

### Risk of Bias Within Studies

After PROBAST assessment had reached consensus, nine (28.1%) studies presented with high AC, whilst all 32 studies demonstrated a high RoB (Table 2). AC for the participant domain were all low, but high for eight studies in the predictor domain, which was related to the inclusion of response to treatment as a prognostic factor. AC was also high for two studies in the outcome domain.^49^ RoB was generally low in the participant domain, but universally high in the predictor, outcome, and analysis domains.

**Table 2.**
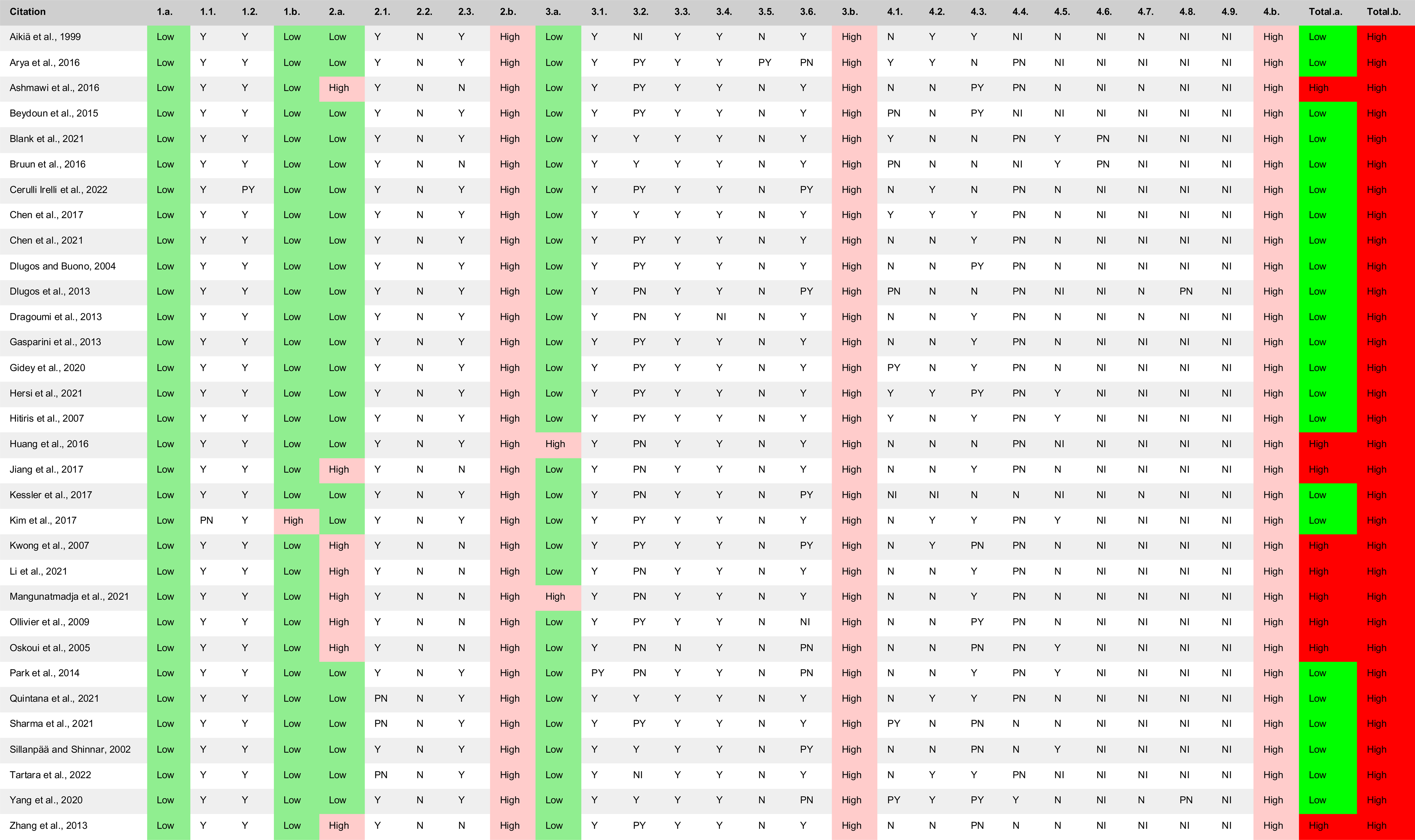
Summary of the PROBAST risk of bias and applicability concern assessment of the included studies. Responses, in order of low to high risk of bias: Y = yes; PY = probably yes; NI = no information; PN = probably no; N = no. x.a. = Domain applicability; x.b. = Domain risk of bias.^16,39,42,61–89^

### Results of Individual Studies

In the included studies outcomes were most commonly categorised as short-term treatment response, followed by seizure remission, pharmacoresistance, and then mortality. Across the multivariable models, 112 relationships between predictors and outcomes were found to be statistically significant, with variables from the semiology category being reported as significant most frequently. Response variables were the next most frequent, followed by history and comorbidity. As shown in table 3, there were 40 cases of variables being statistically significant as predictors of seizure remission, of which 13 were categorised as response variables.

**Table 3.** Summary count of predictor instances, sorted by outcome.

| Predictor Category | Mortality | Pharmacoresistance | Seizure remission | Short-term treatment response | Total |
| --- | --- | --- | --- | --- | --- |
| Age | 1 | 1 | 2 | 5 | 9 |
| ASM | 0 | 1 | 0 | 6 | 7 |
| Comorbidity | 1 | 3 | 1 | 5 | 10 |
| Demographics | 5 | 0 | 1 | 0 | 6 |
| Diagnosis | 1 | 1 | 2 | 2 | 6 |
| EEG | 0 | 2 | 1 | 2 | 5 |
| History | 0 | 0 | 6 | 4 | 10 |
| Neuroimaging | 0 | 0 | 2 | 5 | 7 |
| Neuropsychology | 0 | 0 | 1 | 0 | 1 |
| Response | 0 | 1 | 13 | 2 | 16 |
| Semiology | 1 | 6 | 11 | 17 | 35 |
| Total | 9 | 15 | 40 | 48 | 112 |

**Figure 3.**
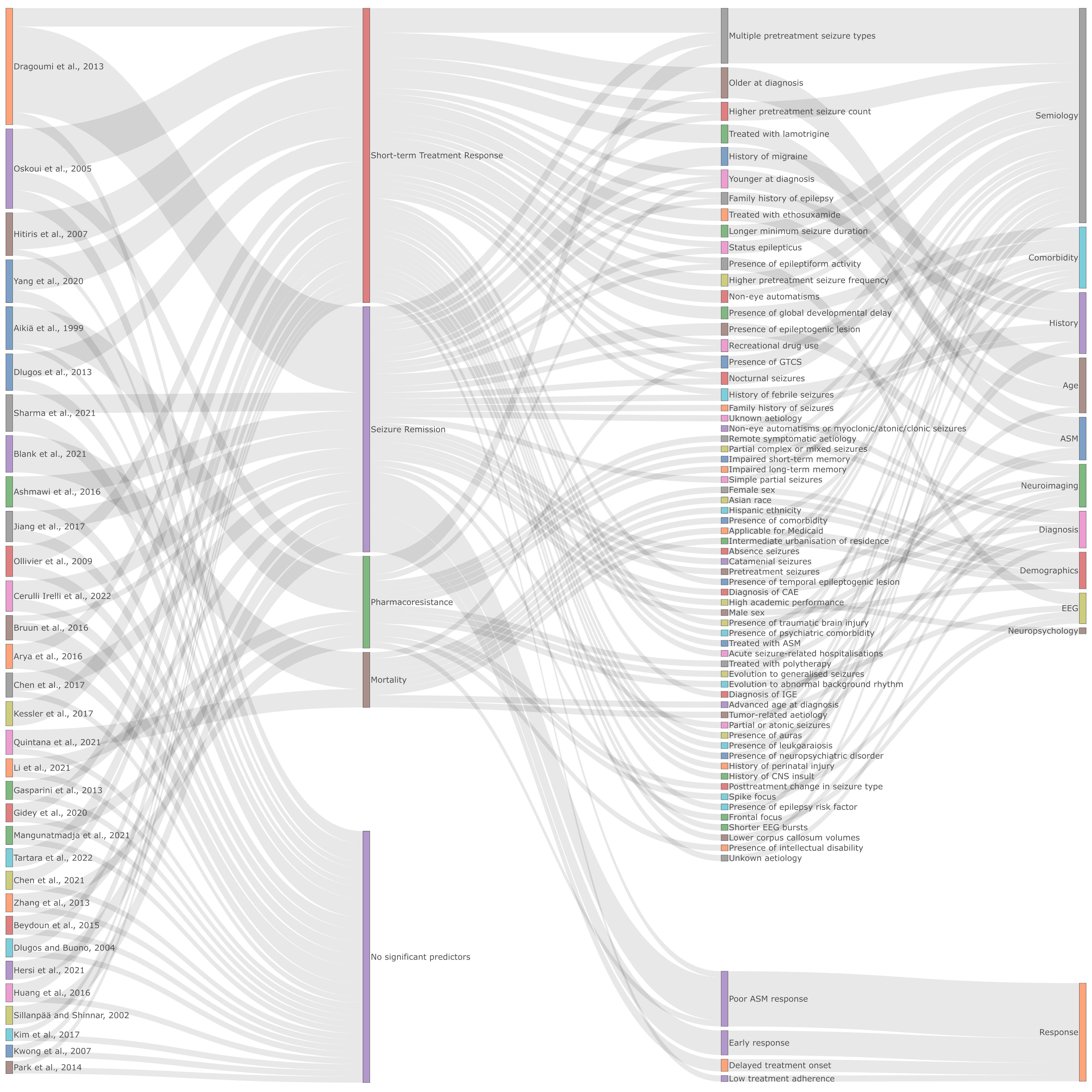
Sankey diagram showing studies and predictors, grouped by outcomes.^16,39,42,61–89^

## Discussion

### Summary of Evidence Review summary

The authors systematically identified 32 studies that used multivariable prediction models to assess the multifactorial prognosis of treatment outcomes in NDE. NDE is an area of research importance in the exploration of the pathomechanisms underlying the development of an epileptogenic environment, and aside from integrating recent research, this review expands on previous NDE prediction model reviews in two ways. First, the most recent review by Abimbola et al. in 2014 did not include studies with sample sizes below 100, which this review does.^50^ Broadening the inclusion criteria for studies facilitates iteration of the review question over time, and encourages the exploration of specific research questions within the same area. Second, quality (in the form of RoB and AC) assessment of the included studies was carried out by two independent reviewers (CR and VP) in accordance with PROBAST, the first review of epilepsy prediction models to do so.^49^ To best meet our primary objective—to inform the prognostic factor choices of future prediction model studies—this report has been prepared in accordance with PRISMA (where appropriate), ensuring maximum transparency, reproducibility, and clarity.^60^

### Model descriptives

The predictors and the outcomes of the included studies were heterogenous, so were stratified into categories to aid interpretation. The most common outcome was short-term treatment response followed by seizure remission, which aligned with our secondary and primary outcomes of interest, respectively. Models of pharmacoresistance and mortality were also reviewed, which address two of the potential treatment failure outcomes. One fifth (20%) of the studies included in this review included treatment response variables as predictors, which limits the applicability of the resultant models. Although statistical significance does not always confer clinical importance, prediction models are at their most informative when being used to inform treatment initiation, i.e. at baseline/diagnosis. Unsurprisingly, treatment response was often a statistically significant prognostic factor of treatment outcome, and potentially obscured predictive relationships that are interrogatable at baseline - such as treatment decision. In the included studies, seizure characteristics and epilepsy history were frequent statistically significant prognostic factors for seizure remission. For research and prediction model purposes, detailed seizure descriptions and family/personal history factors should be collected at baseline. Furthermore, the consistency of prediction models in epilepsy research would be improved by avoiding convenience-based decisions when designing studies, i.e. using a homogenous, predetermined timepoint, instead of the last follow-up. This could be achieved with the development of a Core Outcome Set for NDE.^90,91^

### Bias

The models were found to contain a moderate amount of AC, but universally high RoB. Whilst it seems unlikely that models created in a clinical context can be free of RoB, there are steps that can be taken to minimise it - thus increasing the applicability of any resultant models. For example, adherence to the modelling guidelines of TRIPOD by journal editors (similar to CONSORT) and researchers, and nominal acknowledgement of best practices, such as clearly reporting study and model characteristics, would facilitate research communication and uptake.^92^ Whilst AC were much less common in the included studies, simple steps such as ensuring consistency between study objectives and methods can be taken to help alleviate them in future studies.

### Limitations

Whilst all individuals with new-onset epilepsy have (by definition) NDE, the inverse is not true—some PWE may have an undisclosed or unreported history of seizures, extending beyond the recommended 12 month cut-off.^10^ In consideration of its distinction from NDE, this review has purposefully avoided misattributing any samples as ‘new-onset epilepsy’, instead opting for the more verifiable NDE label. With this omission comes a potential loss of specificity that may hamper the accuracy of the presented model to certain PWE; guidelines for reporting seizure histories have been suggested, which should help to prevent this necessity in future reports.^93^

By including only studies involving a discrete sample of ILAE compliant NDE, this review addresses a sample who are not vulnerable to the common confounds of epilepsy research (such as ASM use and chronicity), or the heterogeneity of broader seizure research.^10^ However, this specificity comes at the cost of generalisability to provoked seizure research. The exploration of febrile, traumatic, and other acute seizure activity also has the potential to elucidate the pathomechanisms of ictogenesis, with ostensible benefit to unprovoked seizure research. Indeed, the two categories of predictors that were the strongest prognostic factors of epileptic seizure remission in this review, history and semiology, allude to pathomechanistic vulnerabilities (respectively, a predisposition to ictogenesis and the seizure insult) that could potentially describe provocation once fully understood. The conclusions of this review should be weighed against those of reviews into early and first seizures of mixed aetiology, to fully understand the influence of precipitation on ictogenesis.^57,94^

Machine learning is a rapidly expanding field in the health data sciences, which has demonstrated widespread potential utility.^58^ Machine learning prediction models were omitted from this review to ensure the comparability of the included regression model studies, and should not be taken as a dismissal of their increasing value to prognosis and diagnosis. Traditional multivariable models have the potential to inform the design of machine learning models, however, further exploration is necessary to evaluate the current state of machine learning prediction modelling in epilepsy. To facilitate the systematic comparison of artificial intelligence prediction model studies, reporting guidelines (TRIPOD-AI and PROBAST-AI) are in development.^95^

### Conclusions

The studies included in this review are heterogenous in both predictor and outcome selection, which is a hindrance to systematic comparison. To evaluate their effectiveness, prediction modelling of treatment decisions should be encouraged, whilst the inclusion of response to treatment as a prognostic factor should be avoided. Authors should also attempt to ensure that their studies adhere to reporting guidelines, to reduce RoB and AC.

## Data Availability

All data produced in the present study are available upon reasonable request to the authors

## Abbreviations

NDE: Newly Diagnosed Epilepsy
PWE: People With Epilepsy
ASM: Anti-seizure Medication(s)
RCT: Randomised Control Trial
RoB: Risk of Bias
AC: Applicability Concerns
TRIPOD(-AI): Transparent Reporting of a multivariable prediction model for Individual Prognosis Or Diagnosis (Artificial Intelligence)
PROBAST(-AI): Prediction model Risk Of Bias ASsessment Tool (Artificial Intelligence)
PRISMA: Preferred Reporting Items for Systematic reviews and Meta-Analyses

## Postscript

### Acknowledgements

Nothing to acknowledge.

### Contributions

CR: Conceptualisation, investigation, visualisation, writing - original draft preparation, writing - review and editing. ORCID ID: https://orcid.org/0000-0002-3824-1681

VP: Investigation.

AM: Writing - review and editing. ORCID ID: https://orcid.org/0000-0002-6861-8806

SSK: Writing - review and editing, supervision. ORCID ID: https://orcid.org/0000-0001-5247-9795

LJB: Conceptualisation, investigation, writing - review and editing. ORCID ID: https://orcid.org/0000-0002-6981-9212

### Ethics and Integrity

Nothing to declare.

