## Appendix 1; Appendix 2; Appendix 3 for "Clinical Prediction Models for Treatment Outcomes in Newly-diagnosed Epilepsy"

### Appendix 1

#### Appendix 1.a.

MEDLINE search strategy (Medical subject headings), carried out on 24.08.22

1. early diagnosis/

2. ((recent$ or new$ or early) adj2 (diagnos$ or onset)).tw.

3. ("first seizure" or "first fit").tw.

4. 1 or 2 or 3

5. exp Epilepsy/ or epilep$.tw.

6. (validation studies or clinical trial or clinical trial phase i or clinical trial phase ii or clinical trial phase iii or clinical trial phase iv or comparative study or evaluation studies or multicenter study).pt.

7. ((observation$ or cohort or case$ or cross?section$ or "cross section$" or "time‐series" or "time series" or "before and after" or "before‐and‐after" or retrospective) adj2 (study or trial or method)).mp.

8. (randomized controlled trial or controlled clinical trial).pt. or (randomized or placebo or randomly).ab.

9. clinical trials as topic.sh.

10. trial.ti.

11. 6 or 7 or 8 or 9 or 10

12. exp animals/ not humans.sh.

13. 11 not 12

14. 13 not case reports.pt.

15. Validat$.mp. or Predict$.ti. or Rule$.mp. or (Predict$ and (Outcome$ or Risk$ or Model$)).mp. or ((History or Variable$ or Criteria or Scor$ or Characteristic$ or Finding$ or Factor$) and (Predict$ or Model$ or Decision$ or Indentif$ or Prognos$)).mp. or (Decision$.mp. and ((Model$ or Clinical$).mp. or Logistic Models/)) or (Prognostic and (History or Variable$ or Criteria or Scor$ or Characteristic$ or Finding$ or Factor$ or Model$)).mp. [mp=title, book title, abstract, original title, name of substance word, subject heading word, floating sub-heading word, keyword heading word, organism supplementary concept word, protocol supplementary concept word, rare disease supplementary concept word, unique identifier, synonyms]

16. 5 and 14 and 15

17. 4 and 16

#### Appendix 1.b.

SCOPUS search strategy (Boolean), carried out on 14.11.22

(
 (
 TITLE-ABS-KEY("early diagnosis")
 OR
 TITLE-ABS-KEY((recent* OR new* OR early*) Pre/0 (diagnos* OR onset))
 OR
 TITLE-ABS-KEY("first seizure" OR "first fit")
 )
 AND
 DOCTYPE(AR)
 AND
 (
 TITLE-ABS-KEY(Epilep*)
 AND
 (
 (
 TITLE-ABS-KEY((observation OR cohort OR case OR "cross section*" OR "time series" OR "before and after" OR retrospective) Pre/0 (study OR trial OR method))
 OR
 ABS("randomized controlled trial" OR "controlled clinical trial" OR randomized OR placebo OR randomly)
 OR
 KEY("clinical trial")
 OR
 TITLE(trial)
 )
 AND NOT
 ALL(animal OR "case report")
 )
 AND
 (
 TITLE-ABS-KEY(validat* OR rule* OR (Predict* AND (Outcome* OR Risk* OR Model*)) OR ((History OR Variable$ OR Criteria OR Scor* OR Characteristic* OR Finding* OR Factor*) AND (Predict* OR Model* OR Decision* OR Indentif* OR Prognos*)) OR (Decision* AND ((Model* OR Clinical*) OR "Logistic Models")) OR (Prognostic AND (History OR Variable* OR Criteria OR Scor* OR Characteristic* OR Finding* OR Factor* OR Model*)))
 OR
 TITLE(predict*)
 )
 )
)

### Appendix 2

#### Predictors

- Age: Factors derived from the age of the patient at diagnosis, or seizure onset. Not the same as disease duration.
- Anti-seizure Medication (ASM): Factors derived from treatment with ASM, such as first-line therapy or ASM change.
- Comorbidity: Factors derived from the presence of a concurrent medical, neuropsychological, or neuropsychiatric condition. Likely operationalised as an ordinal or nominal variable.
- Demographics: Factors derived from the patient’s non-clinical psychosocial environment, excluding age.
- Diagnosis: Factors derived from the patient’s clinical diagnosis with an epilepsy disorder. Likely inferential, based on age, semiology, EEG, neuroimaging, and history.
- Electroencephalography (EEG): Factors derived from functional brain activity data generated with EEG or magnetoencephalography.
- History: Factors derived from the clinical history of the patient and the patient’s family.
- Neuroimaging: Factors derived from any of the following imaging paradigms, in isolation or combination: Magnetic resonance imaging, positron emission tomography, computerised tomography.
- Neuropsychology: Factors derived from sub-clinical neuropsychological performance. Likely operationalised on a continuous scale of performance.
- Response: Factors derived from the patient’s response (disease course) following a medical intervention/treatment plan, i.e. a prescription of ASM.
- Semiology: Factors relating to the dynamics and properties of the seizures experienced by the patient. Not always the same as diagnosis.

#### Outcomes

- Mortality: Endpoints referring to patient death/rate of death, either disease-related or otherwise, at any time point.
- Pharmacoresistance: Endpoints referring to intractability, refractoriness, or poor outcomes in the long-term (>= 12 months).
- Seizure Remission: Endpoints referring to seizure freedom or remission in the long term (>= 12 months).
- Short-term Treatment Response: Endpoints referring to short term (< 12 months) response (disease course) following a medical intervention/treatment plan, i.e. a prescription of ASM.

### Appendix 3

| **Citation** | **Outcome - Verbatim** | **Outcome - Category** | **Outcome - Valence** | **Outcome - Operationalised** | **Predictor - Verbatim** | **Predictor - General** | **Predictor - Category** |
| --- | --- | --- | --- | --- | --- | --- | --- |
| Aikiä et al., 1999 | Refractory seizure disorder | Pharmacoresistance | Positive | Reduced chance of pharmacoresistance | NA | No significant predictors | NA |
| Aikiä et al., 1999 | Refractory seizure disorder | Pharmacoresistance | Negative | Increased chance of pharmacoresistance | Age at diagnosis (Younger = greater likelihood of poor 2-year outcome) | Younger at diagnosis | Age |
| Aikiä et al., 1999 | Refractory seizure disorder | Pharmacoresistance | Negative | Increased chance of pharmacoresistance | Aetiology (Remote symptomatic = greater likelihood of poor 2-year outcome) | Remote symptomatic aetiology | Diagnosis |
| Aikiä et al., 1999 | Refractory seizure disorder | Pharmacoresistance | Negative | Increased chance of pharmacoresistance | Seizure type (Partial complex or mixed = greater likelihood of poor 2-year outcome) | Partial complex or mixed seizures | Semiology |
| Aikiä et al., 1999 | Refractory seizure disorder | Pharmacoresistance | Negative | Increased chance of pharmacoresistance | Spike focus (Presence = greater likelihood of poor 2-year outcome) | Spike focus | EEG |
| Aikiä et al., 1999 | Refractory seizure disorder | Pharmacoresistance | Negative | Increased chance of pharmacoresistance | Immediate list recall (Impairment = greater likelihood of poor 2-year outcome) | Impaired short-term memory | Comorbidity |
| Aikiä et al., 1999 | Refractory seizure disorder | Pharmacoresistance | Negative | Increased chance of pharmacoresistance | Delayed list recognition (Impairment = greater likelihood of poor 2-year outcome) | Impaired long-term memory | Comorbidity |
| Arya et al., 2016 | Freedom from failure | Short-term Treatment Response | Positive | Improved short-term treatment response | NA | No significant predictors | NA |
| Arya et al., 2016 | Freedom from failure | Short-term Treatment Response | Negative | Impaired short-term treatment response | ASM (LTG = reduced chance of freedom from failure) | Treated with lamotrigine | ASM |
| Arya et al., 2016 | Seizure freedom | Short-term Treatment Response | Positive | Improved short-term treatment response | NA | No significant predictors | NA |
| Arya et al., 2016 | Seizure freedom | Short-term Treatment Response | Negative | Impaired short-term treatment response | ASM (LTG = reduced chance of seizure freedom) | Treated with lamotrigine | ASM |
| Ashmawi et al., 2016 | 2-year seizure remission | Seizure Remission | Positive | Increased chance of seizure remission | NA | No significant predictors | NA |
| Ashmawi et al., 2016 | 2-year seizure remission | Seizure Remission | Negative | Reduced chance of seizure remission | Nocturnal seizures (Yes = reduced chance of sustained 2 year remission) | Nocturnal seizures | Semiology |
| Ashmawi et al., 2016 | 2-year seizure remission | Seizure Remission | Negative | Reduced chance of seizure remission | First ASM response (Bad = reduced chance of 2 year remission) | Poor ASM response | Response |
| Ashmawi et al., 2016 | 2-year sustained seizure remission | Seizure Remission | Positive | Increased chance of seizure remission | NA | No significant predictors | NA |
| Ashmawi et al., 2016 | 2-year sustained seizure remission | Seizure Remission | Negative | Reduced chance of seizure remission | First ASM response (Bad = reduced chance of 2 year sustained remission) | Poor ASM response | Response |
| Beydoun et al., 2015 | 6-month terminal seizure remission at month 12 | Seizure Remission | Positive | Increased chance of seizure remission | NA | No significant predictors | NA |
| Beydoun et al., 2015 | 6-month terminal seizure remission at month 12 | Seizure Remission | Negative | Reduced chance of seizure remission | Epileptogenic lesion on neuroimaging (Yes = less likely to experience 6-month terminal remission at month 12) | Presence of epileptogenic lesion | Neuroimaging |
| Beydoun et al., 2015 | 6-month terminal seizure remission at month 12 | Seizure Remission | Negative | Reduced chance of seizure remission | Baseline seizure type (Simple partial = less likely to experience 6-month terminal remission at month 12) | Simple partial seizures | Semiology |
| Blank et al., 2021 | 5-year mortality | Mortality | Positive | Reduced chance of mortality | Sex (Female = decreased risk of mortality) | Female sex | Demographics |
| Blank et al., 2021 | 5-year mortality | Mortality | Positive | Reduced chance of mortality | Race (Asian = decreased risk of mortality) | Asian race | Demographics |
| Blank et al., 2021 | 5-year mortality | Mortality | Positive | Reduced chance of mortality | Ethnicity (Hispanic = decreased risk of mortality) | Hispanic ethnicity | Demographics |
| Blank et al., 2021 | 5-year mortality | Mortality | Negative | Increased chance of mortality | Comorbidity (Yes = increased risk of mortality) | Presence of comorbidity | Comorbidity |
| Blank et al., 2021 | 5-year mortality | Mortality | Negative | Increased chance of mortality | Medicaid coinsurance (Yes = increased risk of mortality) | Applicable for Medicaid | Demographics |
| Blank et al., 2021 | 5-year mortality | Mortality | Negative | Increased chance of mortality | Rural-urban continuum code (Intermediate = increased risk of mortality) | Intermediate urbanisation of residence | Demographics |
| Bruun et al., 2016 | 2-year seizure remission | Seizure Remission | Positive | Increased chance of seizure remission | NA | No significant predictors | NA |
| Bruun et al., 2016 | 2-year seizure remission | Seizure Remission | Negative | Reduced chance of seizure remission | Seizure remission within the first year of ASM treatment (No = less likely to attain 2-year remission) | Poor ASM response | Response |
| Bruun et al., 2016 | 5-year seizure remission | Seizure Remission | Positive | Increased chance of seizure remission | NA | No significant predictors | NA |
| Bruun et al., 2016 | 5-year seizure remission | Seizure Remission | Negative | Reduced chance of seizure remission | NA | No significant predictors | NA |
| Cerulli Irelli et al., 2022 | 4-year seizure remission | Seizure Remission | Positive | Increased chance of seizure remission | NA | No significant predictors | NA |
| Cerulli Irelli et al., 2022 | 4-year seizure remission | Seizure Remission | Negative | Reduced chance of seizure remission | Absence seizures (Present = lower remission probability) | Absence seizures | Semiology |
| Cerulli Irelli et al., 2022 | Delayed seizure remission | Seizure Remission | Positive | Increased chance of seizure remission | NA | No significant predictors | NA |
| Cerulli Irelli et al., 2022 | Delayed seizure remission | Seizure Remission | Negative | Reduced chance of seizure remission | Age at onset (Earlier = remission delay) | Younger at diagnosis | Age |
| Cerulli Irelli et al., 2022 | Delayed seizure remission | Seizure Remission | Negative | Reduced chance of seizure remission | Catamenial seizures (Present = remission delay) | Catamenial seizures | Semiology |
| Chen et al., 2017 | Terminal seizure outcome | Seizure Remission | Positive | Increased chance of seizure remission | NA | No significant predictors | NA |
| Chen et al., 2017 | Terminal seizure outcome | Seizure Remission | Negative | Reduced chance of seizure remission | Seizures in the year prior to treatment (More = poorer chance of seizure freedom) | Pretreatment seizures | Semiology |
| Chen et al., 2017 | Terminal seizure outcome | Seizure Remission | Negative | Reduced chance of seizure remission | Recreational drug use (Yes = poorer chance of seizure freedom) | Recreational drug use | Comorbidity |
| Chen et al., 2017 | Terminal seizure outcome | Seizure Remission | Negative | Reduced chance of seizure remission | Family history of epilepsy (More = poorer chance of seizure freedom) | Family history of epilepsy | History |
| Chen et al., 2021 | Treatment response | Short-term Treatment Response | Negative | Impaired short-term treatment response | Age at onset (Below 5 = lower likelihood of treatment response) | Unknown aetiology | Age |
| Chen et al., 2021 | Treatment response | Short-term Treatment Response | Negative | Impaired short-term treatment response | Attack frequency (Higher = lower likelihood of treatment response) | Higher pretreatment seizure frequency | Semiology |
| Chen et al., 2021 | Treatment response | Short-term Treatment Response | Positive | Improved short-term treatment response | NA | No significant predictors | NA |
| Dlugos and Buono, 2004 | Persistence of LOC seizures at a maximally tolerated dose of CBZ within 1 year of initiation | Short-term Treatment Response | Positive | Improved short-term treatment response | NA | No significant predictors | NA |
| Dlugos and Buono, 2004 | Persistence of LOC seizures at a maximally tolerated dose of CBZ within 1 year of initiation | Short-term Treatment Response | Negative | Impaired short-term treatment response | Early risk factor for epilepsy (Yes = higher chance of trial failure) | Presence of epilepsy risk factor | Neuroimaging |
| Dlugos and Buono, 2004 | Persistence of LOC seizures at a maximally tolerated dose of CBZ within 1 year of initiation | Short-term Treatment Response | Negative | Impaired short-term treatment response | Temporal neuroimaging abnormality (Yes = higher chance of trial failure) | Presence of temporal epileptogenic lesion | Neuroimaging |
| Dlugos et al., 2013 | Freedom from failure at 16 - 20 weeks | Short-term Treatment Response | Positive | Improved short-term treatment response | ASM (ETX over LTG = greater chance of freedom from failure) | Treated with ethosuxamide | ASM |
| Dlugos et al., 2013 | Freedom from failure at 16 - 20 weeks | Short-term Treatment Response | Positive | Improved short-term treatment response | Shortest seizure duration (Longer = greater chance of freedom from failure) | Longer minimum seizure duration | Semiology |
| Dlugos et al., 2013 | Freedom from failure at 16 - 20 weeks | Short-term Treatment Response | Negative | Impaired short-term treatment response | NA | No significant predictors | NA |
| Dlugos et al., 2013 | Seizure freedom at 16 - 20 weeks | Short-term Treatment Response | Positive | Improved short-term treatment response | ASM (ETX over LTG = greater chance of seizure freedom) | Treated with ethosuxamide | ASM |
| Dlugos et al., 2013 | Seizure freedom at 16 - 20 weeks | Short-term Treatment Response | Positive | Improved short-term treatment response | Shortest seizure duration (Longer = greater chance of seizure freedom) | Longer minimum seizure duration | Semiology |
| Dlugos et al., 2013 | Seizure freedom at 16 - 20 weeks | Short-term Treatment Response | Negative | Impaired short-term treatment response | NA | No significant predictors | NA |
| Dragoumi et al., 2013 | 12-month seizure remission at 2 years | Seizure Remission | Positive | Increased chance of seizure remission | Diagnosis (CAE = increased chance of remission at 2 years) | Diagnosis of CAE | Diagnosis |
| Dragoumi et al., 2013 | 12-month seizure remission at 2 years | Seizure Remission | Positive | Increased chance of seizure remission | Response (Early = increased chance of remission at 2 years) | Early response | Response |
| Dragoumi et al., 2013 | 12-month seizure remission at 2 years | Seizure Remission | Negative | Reduced chance of seizure remission | NA | No significant predictors | NA |
| Dragoumi et al., 2013 | Occurrence of seizures in the initial 12 months | Short-term Treatment Response | Positive | Improved short-term treatment response | Age at onset (Older = decreased chance of seizure occurrence in the first 12 months) | Older at diagnosis | Age |
| Dragoumi et al., 2013 | Occurrence of seizures in the initial 12 months | Short-term Treatment Response | Negative | Impaired short-term treatment response | Status epilepticus (Yes = increased chance of seizure occurrence in the first 12 months) | Status epilepticus | Semiology |
| Dragoumi et al., 2013 | Occurrence of seizures in the initial 12 months | Short-term Treatment Response | Negative | Impaired short-term treatment response | Multiple seizure types (More = increased chance of seizure occurrence in the first 12 months) | Multiple pretreatment seizure types | Semiology |
| Dragoumi et al., 2013 | Occurrence of seizures in the preceding 2 years at 4 years | Seizure Remission | Positive | Increased chance of seizure remission | Academic performance (High = decreased chance of seizure occurrence in the preceding 2 years at 4 years) | High academic performance | Neuropsychology |
| Dragoumi et al., 2013 | Occurrence of seizures in the preceding 2 years at 4 years | Seizure Remission | Negative | Reduced chance of seizure remission | History - febrile seizures (Yes = increased chance of seizure occurrence in the preceding 2 years at 4 years) | History of febrile seizures | History |
| Dragoumi et al., 2013 | Occurrence of seizures in the preceding 2 years at 4 years | Seizure Remission | Negative | Reduced chance of seizure remission | History - migraine (Yes = increased chance of seizure occurrence in the preceding 2 years at 4 years) | History of migraine | History |
| Dragoumi et al., 2013 | Occurrence of seizures in the preceding 2 years at study end | Seizure Remission | Positive | Increased chance of seizure remission | NA | No significant predictors | NA |
| Dragoumi et al., 2013 | Occurrence of seizures in the preceding 2 years at study end | Seizure Remission | Negative | Reduced chance of seizure remission | Multiple seizure types (More = increased chance of seizure occurrence in the preceding 2 years at study end) | Multiple pretreatment seizure types | Semiology |
| Dragoumi et al., 2013 | Occurrence of seizures in the preceding 2 years at study end | Seizure Remission | Negative | Reduced chance of seizure remission | Early response (No = increased chance of seizure occurrence in the preceding 2 years at study end) | Poor ASM response | Response |
| Dragoumi et al., 2013 | Occurrence of seizures in the preceding 2 years at study end | Seizure Remission | Negative | Reduced chance of seizure remission | History - migraine (Yes = Increased chance of seizure occurrence in the preceding 2 years at study end) | History of migraine | History |
| Dragoumi et al., 2013 | Occurrence of seizures in the preceding 2 years at study end | Seizure Remission | Negative | Reduced chance of seizure remission | Inital response to treatment (No = increased chance of seizure occurrence in the preceding 2 years at study end) | Poor ASM response | Response |
| Dragoumi et al., 2013 | Remission - relapse pattern | Seizure Remission | Positive | Increased chance of seizure remission | Age at onset (Older = decreased chance pattern 'C') | Older at diagnosis | Age |
| Dragoumi et al., 2013 | Remission - relapse pattern | Seizure Remission | Positive | Increased chance of seizure remission | Response (Early = decreased chance pattern 'C') | Early response | Response |
| Dragoumi et al., 2013 | Remission - relapse pattern | Seizure Remission | Positive | Increased chance of seizure remission | Response (Immediate = decreased chance pattern 'C') | Early response | Response |
| Dragoumi et al., 2013 | Remission - relapse pattern | Seizure Remission | Negative | Reduced chance of seizure remission | Multiple seizure types (More = increased chance of pattern 'C') | Multiple pretreatment seizure types | Semiology |
| Dragoumi et al., 2013 | Remission - relapse pattern | Seizure Remission | Negative | Reduced chance of seizure remission | History - migraine (Yes = increased chance of pattern 'C') | History of migraine | History |
| Gasparini et al., 2013 | 5-year seizure remission | Seizure Remission | Positive | Increased chance of seizure remission | Family history (Epilepsy or febrile seizures = increased chance of remission) | Family history of seizures | History |
| Gasparini et al., 2013 | 5-year seizure remission | Seizure Remission | Positive | Increased chance of seizure remission | Lobe localisation (Front = increased chance of remission) | Frontal focus | Semiology |
| Gasparini et al., 2013 | 5-year seizure remission | Seizure Remission | Negative | Reduced chance of seizure remission | NA | No significant predictors | NA |
| Gidey et al., 2020 | Seizure recurrence | Short-term Treatment Response | Positive | Improved short-term treatment response | NA | No significant predictors | NA |
| Gidey et al., 2020 | Seizure recurrence | Short-term Treatment Response | Negative | Impaired short-term treatment response | Pretreatment seizure number (Greater = decreased chance of achieving seizure remission) | Higher pretreatment seizure count | Semiology |
| Gidey et al., 2020 | Seizure recurrence | Short-term Treatment Response | Negative | Impaired short-term treatment response | Treatment adherence (Poor = decreased chance of achieving seizure remission) | Low treatment adherence | Response |
| Hersi et al., 2021 | 12-month seizure remission | Seizure Remission | Positive | Increased chance of seizure remission | Sex (Male = more likely to achieve remission) | Male sex | Demographics |
| Hersi et al., 2021 | 12-month seizure remission | Seizure Remission | Positive | Increased chance of seizure remission | Aetiology (Unknown = more likely to achieve remission) | Unknown aetiology | Diagnosis |
| Hersi et al., 2021 | 12-month seizure remission | Seizure Remission | Negative | Reduced chance of seizure remission | EEG (Epileptiform acivity = less likely to achieve seizure freedom) | Presence of epileptiform activity | EEG |
| Hitiris et al., 2007 | Seizure free for the last 12 months | Short-term Treatment Response | Positive | Improved short-term treatment response | NA | No significant predictors | NA |
| Hitiris et al., 2007 | Seizure free for the last 12 months | Short-term Treatment Response | Negative | Impaired short-term treatment response | Family history - epilepsy (Yes = greater risk of pharmacoresistance) | Family history of epilepsy | History |
| Hitiris et al., 2007 | Seizure free for the last 12 months | Short-term Treatment Response | Negative | Impaired short-term treatment response | History - febrile seizures (Yes = greater risk of pharmacoresistance) | History of febrile seizures | History |
| Hitiris et al., 2007 | Seizure free for the last 12 months | Short-term Treatment Response | Negative | Impaired short-term treatment response | Traumatic brain injury (Yes = greater risk of pharmacoresistance) | Presence of traumatic brain injury | Neuroimaging |
| Hitiris et al., 2007 | Seizure free for the last 12 months | Short-term Treatment Response | Negative | Impaired short-term treatment response | Psychiatric comorbidity (Yes = greater risk of pharmacoresistance) | Presence of psychiatric comorbidity | Comorbidity |
| Hitiris et al., 2007 | Seizure free for the last 12 months | Short-term Treatment Response | Negative | Impaired short-term treatment response | Recreational drug use (Yes = greater risk of pharmacoresistance) | Recreational drug use | Comorbidity |
| Hitiris et al., 2007 | Seizure free for the last 12 months | Short-term Treatment Response | Negative | Impaired short-term treatment response | More than 10 seizures before treatment (Yes = greater risk of pharmacoresistance) | Higher pretreatment seizure count | Semiology |
| Huang et al., 2016 | 50% seizure reduction | Short-term Treatment Response | Positive | Improved short-term treatment response | Age at onset (Older = more likely to achieve satisfactory seizure control at 2 years) | Older at diagnosis | Age |
| Huang et al., 2016 | 50% seizure reduction | Short-term Treatment Response | Positive | Improved short-term treatment response | ASM treatment (Yes = more likely to achieve satisfactory seizure control at 2 years) | Treated with ASM | ASM |
| Huang et al., 2016 | 50% seizure reduction | Short-term Treatment Response | Negative | Impaired short-term treatment response | NA | No significant predictors | NA |
| Jiang et al., 2017 | 2-year seizure remission at short-term follow-up | Seizure Remission | Positive | Increased chance of seizure remission | NA | No significant predictors | NA |
| Jiang et al., 2017 | 2-year seizure remission at short-term follow-up | Seizure Remission | Negative | Reduced chance of seizure remission | Time to treatment (> 12 months = more likely to experience unfavourable short-term outcomes) | Delayed treatment onset | Response |
| Jiang et al., 2017 | 2-year seizure remission at short-term follow-up | Seizure Remission | Negative | Reduced chance of seizure remission | Seizure frequency in the first year of treatment (> 2 = more likely to experience unfavourable short-term outcomes) | Poor ASM response | Response |
| Jiang et al., 2017 | 5-year seizure remission at long-term follow-up | Seizure Remission | Positive | Increased chance of seizure remission | NA | No significant predictors | NA |
| Jiang et al., 2017 | 5-year seizure remission at long-term follow-up | Seizure Remission | Negative | Reduced chance of seizure remission | Seizure frequency in the first year of treatment (> 2 = more likely to experience unfavourable long-term outcomes) | Poor ASM response | Response |
| Kessler et al., 2017 | Seizure freedom at 16 - 20 weeks | Short-term Treatment Response | Positive | Improved short-term treatment response | Shortest burst duration on baseline EEG (Short = higher chance of seizure freedom) | Shorter EEG bursts | EEG |
| Kessler et al., 2017 | Seizure freedom at 16 - 20 weeks | Short-term Treatment Response | Negative | Impaired short-term treatment response | Lamotrigine vs Ethosuximide (Lamotrigine = lower chance of seizure freedom) | Treated with lamotrigine | ASM |
| Kessler et al., 2017 | Seizure freedom at 16 - 20 weeks | Short-term Treatment Response | Negative | Impaired short-term treatment response | Cluster pattern 2 (Yes = lower chance of seizure freedom) | Non-eye automatisms | Semiology |
| Kessler et al., 2017 | Seizure freedom at 16 - 20 weeks | Short-term Treatment Response | Negative | Impaired short-term treatment response | Cluster pattern 2/4 (Yes = lower chance of seizure freedom) | Non-eye automatisms or myoclonic/atonic/clonic seizures | Semiology |
| Kim et al., 2017 | < 6 months of continuous seizure freedom | Short-term Treatment Response | Positive | Improved short-term treatment response | Corpus callosum volume (Lower = good ASM response) | Lower corpus callosum volumes | Neuroimaging |
| Kim et al., 2017 | < 6 months of continuous seizure freedom | Short-term Treatment Response | Negative | Impaired short-term treatment response | NA | No significant predictors | NA |
| Kwong et al., 2007 | Seizure freedom | Short-term Treatment Response | Positive | Improved short-term treatment response | NA | No significant predictors | NA |
| Kwong et al., 2007 | Seizure freedom | Short-term Treatment Response | Negative | Impaired short-term treatment response | Acute seizure-related hospitalisations (Yes = more likely not to acheive seizure-freedom) | Acute seizure-related hospitalisations | Semiology |
| Li et al., 2021 | 3-year seizure freedom | Pharmacoresistance | Positive | Reduced chance of pharmacoresistance | NA | No significant predictors | NA |
| Li et al., 2021 | 3-year seizure freedom | Pharmacoresistance | Negative | Increased chance of pharmacoresistance | Seizure types (Multiple = greater chance of poor drug response) | Multiple pretreatment seizure types | Semiology |
| Li et al., 2021 | 3-year seizure freedom | Pharmacoresistance | Negative | Increased chance of pharmacoresistance | Polytherapy (Yes = greater chance of poor drug response) | Treated with polytherapy | ASM |
| Mangunatmadja et al., 2021 | Intractable epilepsy | Pharmacoresistance | Positive | Reduced chance of pharmacoresistance | NA | No significant predictors | NA |
| Mangunatmadja et al., 2021 | Intractable epilepsy | Pharmacoresistance | Negative | Increased chance of pharmacoresistance | Seizure-type evolution (Generalisation at study end = greater chance of intractability) | Evolution to generalised seizures | Semiology |
| Mangunatmadja et al., 2021 | Intractable epilepsy | Pharmacoresistance | Negative | Increased chance of pharmacoresistance | Background rhythm evolution (Abnormal at study end = greater chance of intractability) | Evolution to abnormal background rhythm | EEG |
| Ollivier et al., 2009 | Complete disappearance of absence seizures during VPA treatment | Short-term Treatment Response | Positive | Improved short-term treatment response | Age at diagnosis (Older = protective factor against non-responsiveness to VPA) | Older at diagnosis | Age |
| Ollivier et al., 2009 | Complete disappearance of absence seizures during VPA treatment | Short-term Treatment Response | Negative | Impaired short-term treatment response | Pretreatment seizure frequency > 10/day (Yes = risk factor for non-responsiveness to VPA) | Higher pretreatment seizure frequency | Semiology |
| Ollivier et al., 2009 | Complete disappearance of absence seizures during VPA treatment | Short-term Treatment Response | Negative | Impaired short-term treatment response | Presence of GTCS (Yes = risk factor for non-responsiveness to VPA) | Presence of GTCS | Semiology |
| Ollivier et al., 2009 | Long-term seizure freedom | Seizure Remission | Positive | Increased chance of seizure remission | NA | No significant predictors | NA |
| Ollivier et al., 2009 | Long-term seizure freedom | Seizure Remission | Negative | Reduced chance of seizure remission | NA | No significant predictors | NA |
| Oskoui et al., 2005 | Lower probability of seizure remission at 12 months | Pharmacoresistance | Positive | Reduced chance of pharmacoresistance | NA | No significant predictors | NA |
| Oskoui et al., 2005 | Lower probability of seizure remission at 12 months | Pharmacoresistance | Negative | Increased chance of pharmacoresistance | More than one seizure type (Yes = increased chance of intractability at 12 months) | Multiple pretreatment seizure types | Semiology |
| Oskoui et al., 2005 | Lower probability of seizure remission at 12 months | Pharmacoresistance | Negative | Increased chance of pharmacoresistance | Seizure recurrence in the 6-12 months post treatment (Yes = increased chance of intractability at 12 months) | Poor ASM response | Response |
| Oskoui et al., 2005 | Lower probability of seizure remission at 12 months | Pharmacoresistance | Negative | Increased chance of pharmacoresistance | Mental retardation (Yes = increased chance of intractability at 12 months) | Presence of intellectual disability | Comorbidity |
| Oskoui et al., 2005 | Lower probability of seizure remission at 3 months | Short-term Treatment Response | Positive | Improved short-term treatment response | NA | No significant predictors | NA |
| Oskoui et al., 2005 | Lower probability of seizure remission at 3 months | Short-term Treatment Response | Negative | Impaired short-term treatment response | More than one seizure type (Yes = increased chance of intractability at 3 months) | Multiple pretreatment seizure types | Semiology |
| Oskoui et al., 2005 | Poor outcome at 12 months | Short-term Treatment Response | Positive | Improved short-term treatment response | NA | No significant predictors | NA |
| Oskoui et al., 2005 | Poor outcome at 12 months | Short-term Treatment Response | Negative | Impaired short-term treatment response | More than one seizure type (Yes = increased chance of poor outcome at 12 months) | Multiple pretreatment seizure types | Semiology |
| Oskoui et al., 2005 | Poor outcome at 12 months | Short-term Treatment Response | Negative | Impaired short-term treatment response | Global developmental delay at onset (Yes = increased chance of poor outcome at 12 months) | Presence of global developmental delay | Comorbidity |
| Oskoui et al., 2005 | Poor outcome at 12 months | Short-term Treatment Response | Negative | Impaired short-term treatment response | Seizure recurrence in the 6-12 months post treatment (Yes = increased chance of poor outcome at 12 months) | Poor ASM response | Response |
| Oskoui et al., 2005 | Poor outcome at 3 months | Short-term Treatment Response | Positive | Improved short-term treatment response | Diagnosis (IGE = decreased chance of poor outcome at 3 months) | Diagnosis of IGE | Diagnosis |
| Oskoui et al., 2005 | Poor outcome at 3 months | Short-term Treatment Response | Negative | Impaired short-term treatment response | More than one seizure type (Yes = increased chance of poor outcome) | Multiple pretreatment seizure types | Semiology |
| Oskoui et al., 2005 | Poor outcome at 3 months | Short-term Treatment Response | Negative | Impaired short-term treatment response | Global developmental delay at onset (Yes = increased chance of poor outcome at 3 months) | Presence of global developmental delay | Comorbidity |
| Park et al., 2014 | Seizure free for the last 6 months | Short-term Treatment Response | Positive | Improved short-term treatment response | Age at onset (16+ = increased chance of being a responder) | Older at diagnosis | Age |
| Park et al., 2014 | Seizure free for the last 6 months | Short-term Treatment Response | Negative | Impaired short-term treatment response | NA | No significant predictors | NA |
| Quintana et al., 2021 | Mortality | Mortality | Positive | Reduced chance of mortality | NA | No significant predictors | NA |
| Quintana et al., 2021 | Mortality | Mortality | Negative | Increased chance of mortality | Older age (Higher = increased risk of mortality) | Advanced age at diagnosis | Age |
| Quintana et al., 2021 | Mortality | Mortality | Negative | Increased chance of mortality | Tumor-related etiology (Yes = increased risk of mortality) | Tumor-related aetiology | Diagnosis |
| Quintana et al., 2021 | Mortality | Mortality | Negative | Increased chance of mortality | Generalized seizures (Yes = increased risk of mortality) | Presence of GTCS | Semiology |
| Sharma et al., 2021 | 12-month seizure remission | Seizure Remission | Positive | Increased chance of seizure remission | NA | No significant predictors | NA |
| Sharma et al., 2021 | 12-month seizure remission | Seizure Remission | Negative | Reduced chance of seizure remission | NA | No significant predictors | NA |
| Sharma et al., 2021 | Seizure recurrence | Seizure Remission | Positive | Increased chance of seizure remission | NA | No significant predictors | NA |
| Sharma et al., 2021 | Seizure recurrence | Seizure Remission | Negative | Reduced chance of seizure remission | Epileptogenic neuroimaging findings (Yes = higher rate of seizure recurrence) | Presence of epileptogenic lesion | Neuroimaging |
| Sharma et al., 2021 | Seizure recurrence | Seizure Remission | Negative | Reduced chance of seizure remission | Prediagnosis seizure number (5+ = higher rate of seizure recurrence) | Higher pretreatment seizure count | Semiology |
| Sharma et al., 2021 | Seizure recurrence | Seizure Remission | Negative | Reduced chance of seizure remission | Treatment approach (Deferred = higher rate of seizure recurrence) | Delayed treatment onset | Response |
| Sillanpää and Shinnar, 2002 | 5-year terminal seizure remission | Seizure Remission | Positive | Increased chance of seizure remission | Response (Early = increased probability of remission) | Early response | Response |
| Sillanpää and Shinnar, 2002 | 5-year terminal seizure remission | Seizure Remission | Negative | Reduced chance of seizure remission | Seizure type (Partial or atonic = decreased probability of remission) | Partial or atonic seizures | Semiology |
| Sillanpää and Shinnar, 2002 | 5-year terminal seizure remission | Seizure Remission | Negative | Reduced chance of seizure remission | Status epilepticus (Occurrence = lower rate of remission) | Status epilepticus | Semiology |
| Tartara et al., 2022 | Seizure freedom | Short-term Treatment Response | Positive | Improved short-term treatment response | Aetiology (Unknown = lower risk of recurrence) | Unkown aetiology | Diagnosis |
| Tartara et al., 2022 | Seizure freedom | Short-term Treatment Response | Negative | Impaired short-term treatment response | Subjective perceptions at seizure onset (Presence = higher risk of recurrence) | Presence of auras | Semiology |
| Tartara et al., 2022 | Seizure freedom | Short-term Treatment Response | Negative | Impaired short-term treatment response | Leukoaraiosis (Presence = higher risk of recurrence) | Presence of leukoaraiosis | Neuroimaging |
| Yang et al., 2020 | Seizure freedom at 12 months | Short-term Treatment Response | Positive | Improved short-term treatment response | NA | No significant predictors | NA |
| Yang et al., 2020 | Seizure freedom at 12 months | Short-term Treatment Response | Negative | Impaired short-term treatment response | Circadian rhythm (Seizures in wake and sleep = poor probability of seizure freedom) | Nocturnal seizures | Semiology |
| Yang et al., 2020 | Seizure freedom at 12 months | Short-term Treatment Response | Negative | Impaired short-term treatment response | Pre-ASM EEG (Epileptiform discharges = poor probability of seizure freedom) | Presence of epileptiform activity | EEG |
| Yang et al., 2020 | Seizure freedom at 12 months | Short-term Treatment Response | Negative | Impaired short-term treatment response | Neuropsychiatric disorder (Presence of any = poor probability of seizure freedom) | Presence of neuropsychiatric disorder | Comorbidity |
| Yang et al., 2020 | Seizure freedom at 12 months | Short-term Treatment Response | Negative | Impaired short-term treatment response | Perinatal brain injury (Yes = poor probability of seizure freedom) | History of perinatal injury | History |
| Yang et al., 2020 | Seizure freedom at 12 months | Short-term Treatment Response | Negative | Impaired short-term treatment response | History - CNS infection (Yes = poor probability of seizure freedom) | History of CNS insult | History |
| Yang et al., 2020 | Seizure freedom at 6 months | Short-term Treatment Response | Positive | Improved short-term treatment response | NA | No significant predictors | NA |
| Zhang et al., 2013 | Poor outcome | Pharmacoresistance | Positive | Reduced chance of pharmacoresistance | NA | No significant predictors | NA |
| Zhang et al., 2013 | Poor outcome | Pharmacoresistance | Negative | Increased chance of pharmacoresistance | Multiple seizure type (Yes = greater chance of poor outcome) | Multiple pretreatment seizure types | Semiology |
| Zhang et al., 2013 | Poor outcome | Pharmacoresistance | Negative | Increased chance of pharmacoresistance | Changes in seizure type during treatment (Yes = greater chance of poor outcome) | Posttreatment change in seizure type | Semiology |

Detailed study demographics
